# Social interaction perception in adult-onset craniopharyngioma

**DOI:** 10.1101/2025.09.14.25335715

**Authors:** Ellie Dadds, Katie Daughters

**Author notes:** Corresponding author: Dr Katie Daughters.

## Abstract

Adult-onset craniopharyngioma (AoC) is a rare intracranial tumour associated with long-term physical and psychological difficulties. Although prior questionnaire-based studies suggest impairments in social and emotional functioning, no research has experimentally examined social interaction perception in this population. This study employed an online experimental task combined with automated linguistic analysis to explore differences in social interaction perception between AoC patients and healthy volunteers (HVs). Nineteen AoC patients and twenty matched HVs viewed 30 short, naturalistic video clips depicting everyday social interactions. After each video, participants were asked to describe what they saw in as much detail as possible. Responses were analysed using Linguistic Inquiry and Word Count (LIWC) software to assess emotional and social language use across eight pre-registered categories. AoC participants used a higher proportion of social and emotional language across six categories (e.g., affect, social behaviour) despite producing shorter responses overall. Inferential statistics found no significant differences between groups, and Bayesian analysis confirmed there were no differences in the use of pro-social or emotion words between AoC participants and HVs. Contrary to hypotheses, AoC patients did not significantly differ from HVs in their linguistic descriptions of social interactions. This may indicate preserved social interaction perception or may reflect task or sample limitations. Future studies should explore the influence of hypothalamic involvement and oxytocin in social functioning in AoC.

## Introduction

Adult-onset craniopharyngioma (AoC), a rare benign brain tumour of the sellar and parasellar region of the brain with an incidence of 0.5 to 2 cases per million a year, accounts for 1-3% of primary brain tumours in adults (1,2). It is associated with debilitating life-long physical morbidities and a poorer quality of life (1,3,4). While a few medical studies suggest psychological factors may contribute to this reduced quality of life (5–10), these studies are limited by the use of medical data for inferences or medical questionnaires that are not as detailed as their psychological counterparts. Moreover, questionnaires in general are susceptible to various biases. Thus, more experimental data is desperately needed to further understand the psychological impact of AoC, with a view to identifying potential targets for future interventions to improve patient quality of life.

Three studies have utilised psychological *questionnaires* to assess various aspects of psychological functioning and mental health in AoC patients (11–13). Gebert et al (11) found that, compared to healthy volunteers (HVs), AoC patients reported higher levels of depression and anxiety, but no differences in a measure of their emotional skills. These results were replicated across two studies by the authors (12,13); finding that AoC patients reported higher levels of depression and anxiety compared to HVs, and no difference in a measure of emotion regulation (12); and that AoC patients reported feeling low mood and anxiety but felt they were good at recognising both their own and other’s emotions (13). Crucially, however, both studies also identified additional areas of difficulty. AoC patients scored lower in a measure of social functioning; higher in alexithymia (the ability to detect and identify one’s own emotions); and lower overall life satisfaction (12); as well as reporting missing social events and a shrinking social network (13).

Three studies have employed *experimental* designs to investigate emotional processing in AoC patients (14–16). Across two tasks, participants were shown photographs of facial expressions and asked to either identify the emotion (emotion recognition task) or select a word that best matched the mental state of the individual (theory-of-mind (ToM) task). All three studies found AoC patients performed worse on emotion recognition tasks compared to HVs (14–16). For ToM, one study found AoC patients performed worse (15); one found patients high in apathy performed worse compared to those without apathy (16); and one found no differences (14). For those that collected additional questionnaire data, two found that AoC patients reported higher levels of autistic traits (14,15), one found AoC patients reported lower levels of trait empathy (15), and one found AoC patients reported lower levels of pleasure in social interactions (14).

Taken together, this small body of research indicates a clear psychological impact of AoC. While experimental studies have focused on quantifying emotional processing difficulties, questionnaire data suggest that AoC patients also struggle with social interaction. The aim of the current study, therefore, was to use an experimental design to quantify, for the first time, a measure of social functioning in AoC patients. Specifically, the study explored if social interaction perception is altered in patients with AoC. Based on previous research, it was hypothesised that patients with AoC would rate videos of social interactions as significantly less social and emotional compared to matched HVs.

## Materials and Methods

### Participants

AoC patients were recruited via an existing participant database as well as adverts hosted on social media. In order to take part in the research all AoC participants were required to provide evidence of an AoC diagnosis. When satisfactory evidence was provided, participants were sent a link to take part in the experiment. HVs were recruited via the platform Prolific (https://www.prolific.com/) and were matched for age, gender and nationality to AoC participants. HVs were asked to confirm they had no existing endocrine condition. Recruitment took place between 1^st^ January and 1^st^ December 2024.

The study link was accessed 59 times, and responses were screened in accordance with standard online testing practices and the pre-registration to ensure data quality. Responses without complete informed consent (n=8); responses from an email address that did not have a confirmed AOC diagnosis/did not identify as having AOC/did identify as having an endocrine condition (n=2); responses identified as ‘spam’ by Pavlovia (n=1); responses that took under five minutes (n=0); replicate responses from the same IP address (n=0); responses that completed less than 50% of the experiment (n=9); and finally, responses with a high percentage of repetitive responses to each video clip (n=0) were removed from the dataset. Ultimately, 19 AoC participants (including one childhood-onset craniopharyngioma patient) and 20 HVs took part in the study. Tests confirmed there was no significant difference in sex distribution (*χ*^*2*^(1, *N* = 39) = 0.33, *p* = .855) or age (*t*(37) = 1.68, *p* = .101) across groups. A summary of the sample characteristics is provided in Table 1. The study was approved by the Ethics Committee at the University of Essex and was pre-registered (https://aspredicted.org/qyq6-bd8v.pdf). The data and code are available online (https://osf.io/ux2kp/?view_only=47b518305d0e4cceb156f95a14f6ae80).

**Table 1.**
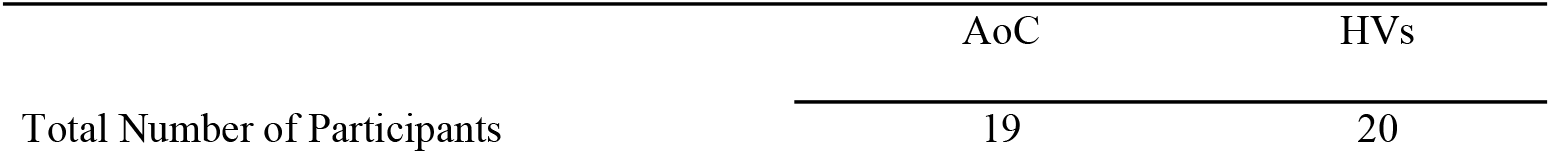
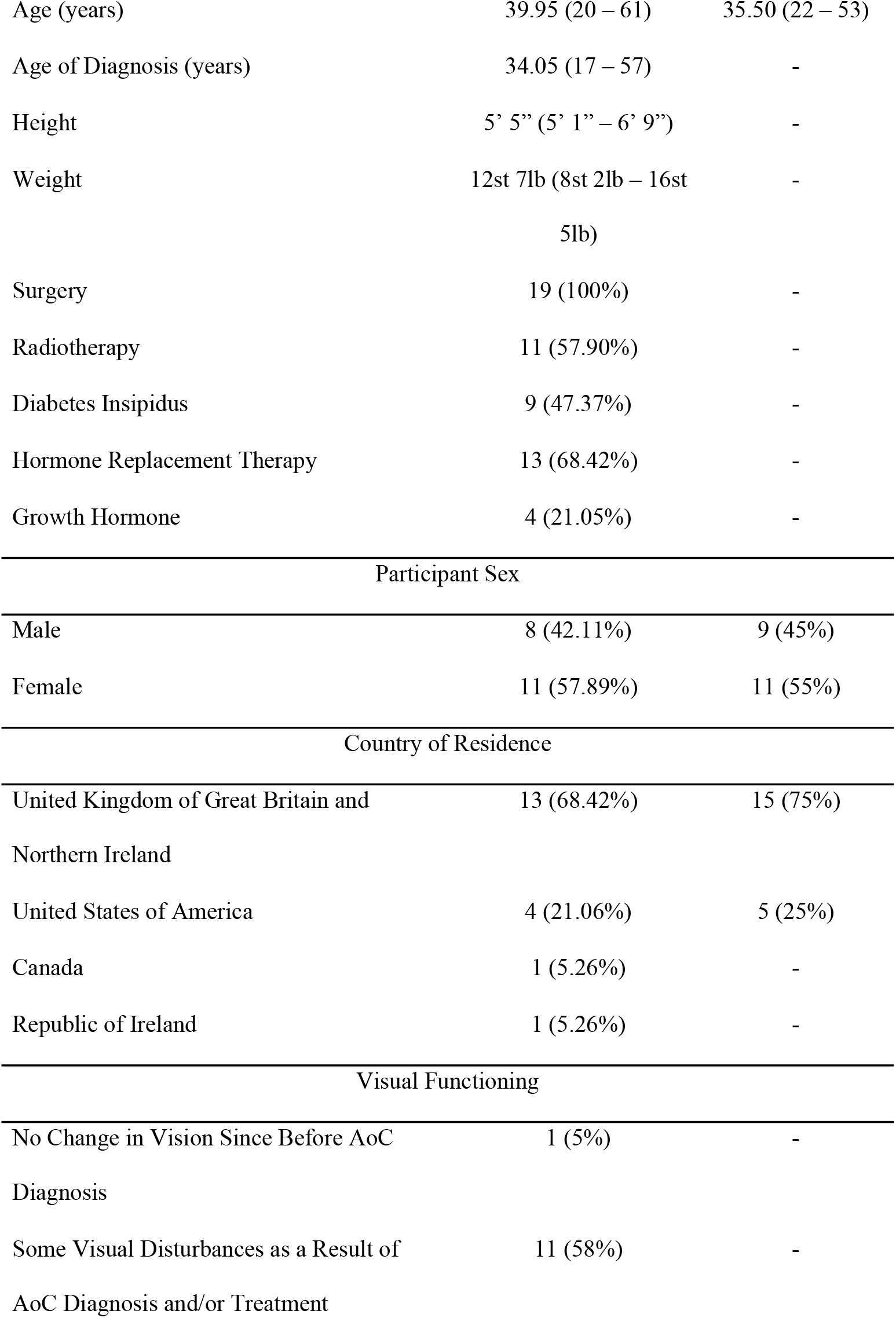
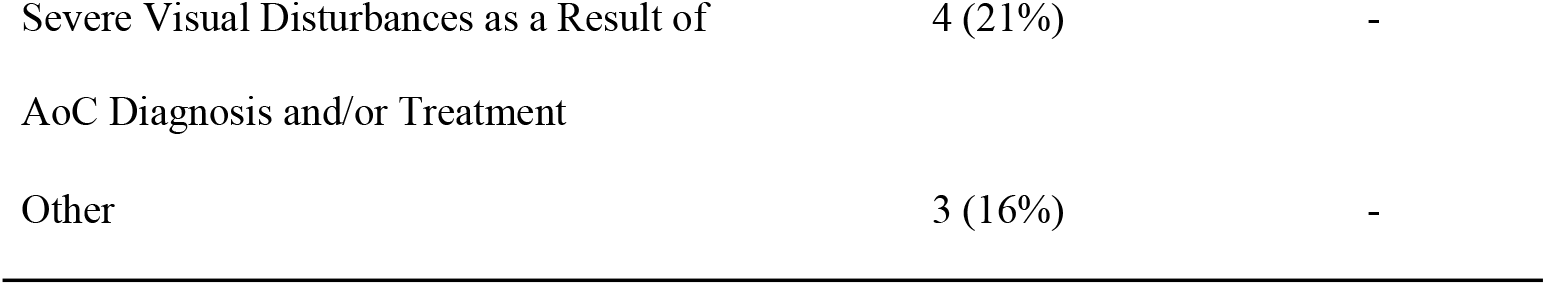
Sample characteristics.

### Materials

The study utilised stimuli that were created and validated for a previous study (17) and have been used to investigate social interaction perception in HVs (18). The stimuli were 60 4-second video clips, showing various everyday scenarios. Each scenario depicted either a social interaction or non-interaction (acting independently) between two individuals. Fifty percent of the video clips showed a social interaction and 50% showed a non-interaction. The video clips of social interactions and non-interactions were matched as carefully as possible, including the same action, props, proximity and physical motion. Scenarios were performed across eight different geographical locations, by four different actor pairings (two female/female pairs; one male/male pair; one female/male pair). The videos did not have audio to avoid confounding variables. Full details are provided in Landsiedel et al. (17) and examples of the stimuli can find online (https://osf.io/cxu9g/?view_only=1cf65251c1c44a26a6a2fbef10cea088). The experiment was created in PsychoPy 3 (19) and presented online via Pavlovia (https://pavlovia.org/).

### Procedure

Participants provided basic demographic information to confirm eligibility before reading the study information sheet and providing written informed consent. Participants read the study instructions and were then shown one video at a time. After each video, they were asked to describe what they thought was happening in as much detail as possible in the free text box provided. Participants were offered a break in the middle of the task. At the end of the study, participants were debriefed and provided £10 ($10) compensation.

### Data Analysis

Participant responses were analysed using Linguistic Inquiry and Word Count (LIWC) to analyse the written content for psychologically meaningful categories (20). LIWC has previously been used to understand language style and how this connects with social relationships and emotionality (21). To do this, LIWC software uses over 100 built-in dictionaries to capture social and psychological states. LIWC calculates the number of words related to each category and provides a percentage. A higher percentage indicates higher use of the words in that specific category. Only responses to the social interaction videos were analysed for the study, as it was not expected that there would be group differences in response to non-interactions.

In line with the pre-registration, 15 relevant categories were identified for analysis: (a) affect; (b) angry emotion; (c) anxious emotion; (d) communication; (e) emotion; (f) feeling; (g) negative emotion; (h) negative tone; (i) positive emotion; (j) positive tone; (k) pro-social behaviour; (l) sad emotion; (m) social; (n) social behaviour; (o) tone. However, because LIWC can only generate scores for categories where the necessary words have been used in a response, for some videos several categories did not generate relevant data. A 50% cut-off was used, such that data had to be present for at least 15 of the 30 videos in order to be included in the analysis. This resulted in six categories (negative tone; negative emotion; anxious emotion; angry emotion; sad emotion; and feeling) - and their corresponding categories (positive tone and positive emotion) - being excluded from analysis. See Table 2 for the eight categories included and exemplar words in each dictionary.

**Table 2.** Exemplar words from LIWC for each category.

| LIWC Category | Exemplar Words |
| --- | --- |
| Affect | Good, well, new |
| Communication | Said, say, tell |
| Emotion | Love, happy, hope |
| Pro-social Behaviour | Care, help, please |
| Social | You, we, he, she |
| Social Behaviour | Love, say, care |
| Tone | Degree of positive/negative tone |
| Word count | Total Word Count |

The data were not normally distributed; Mann Whiney-U tests were used to determine whether AoC participants used statistically different descriptions of social interactions compared to HVs. In light of non-significant findings, exploratory Bayesian analyses were carried out to investigate whether there was greater support for the null hypothesis relative to the alternative hypothesis (22). Data analysis was carried out in IBM SPSS Statistics (Version 29).

## Results

AoC participants scored higher in word use percentages across six of the eight categories: affect, communication, pro-social, social, social behaviour and tone (see Table 3). Interestingly, they did this despite using fewer words compared to HVs. Pre-registered inferential statistical analysis revealed, however, that there were no significant differences in any category in the number of words used to describe social interactions between AoC participants and HVs (see Table 4). To evaluate whether there was greater evidence of the null versus experimental hypothesis Bayes Factors (BF) were calculated using a non-informative prior distribution (see Table 4). The established interpretation of BF suggests that BF > 3 provides moderate evidence that the data support the null hypothesis, and that BF close to 1 suggests data insensitivity (22–24). Following this interpretation, there was moderate evidence for the absence of an effect between groups for the emotion and pro-social categories; in other words, there was over three times more evidence in favour of the null hypothesis. The BFs for the remaining categories suggest data insensitivity, such that there is not enough evidence either way.

**Table 3.**
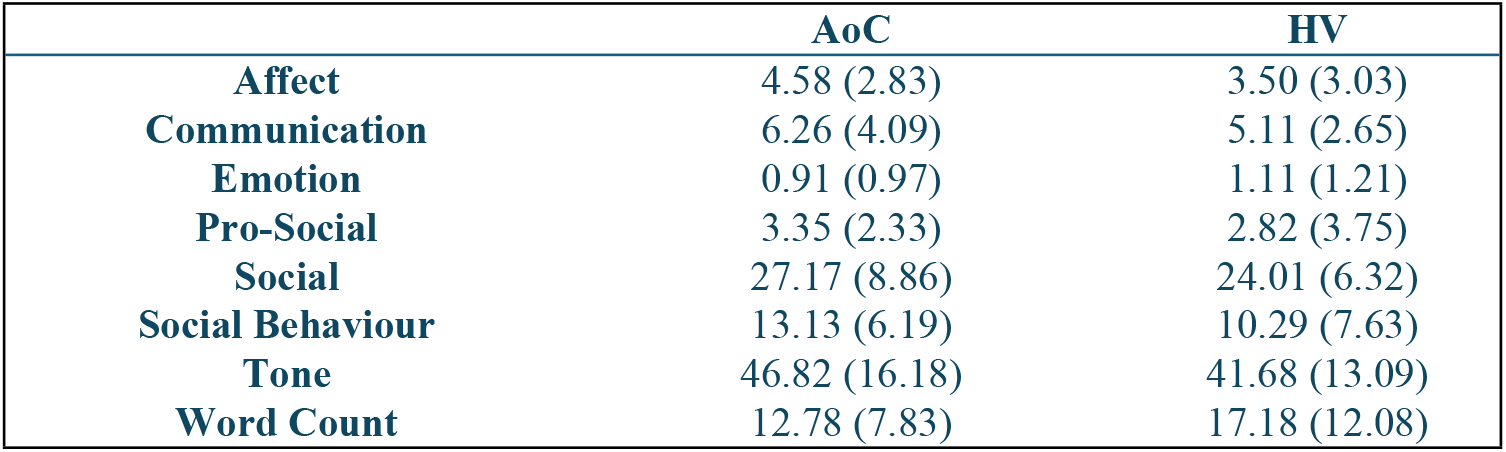
Mean and standard deviation of LIWC percentage per category by group.

|  | **AoC** | **HV** |
| --- | --- | --- |
| **Affect** | 4.58 (2.83) | 3.50 (3.03) |
| **Communication** | 6.26 (4.09) | 5.11 (2.65) |
| **Emotion** | 0.91 (0.97) | 1.11 (1.21) |
| **Pro-Social** | 3.35 (2.33) | 2.82 (3.75) |
| **Social** | 27.17 (8.86) | 24.01 (6.32) |
| **Social Behaviour** | 13.13 (6.19) | 10.29 (7.63) |
| **Tone** | 46.82 (16.18) | 41.68 (13.09) |
| **Word Count** | 12.78 (7.83) | 17.18 (12.08) |

**Table 4.** Mann Whitney U statistics for each LIWC category.

| | $BF_{01}$ | $U$ | $p$ | **MR** | |
| --- | --- | --- | --- | --- | --- |
|  |  |  |  | **AoC** | **HV** |
| **Affect** | 2.397 | 150.00 | .270 | 22.11 | 18.00 |
| **Communication** | 2.642 | 173.00 | .647 | 20.89 | 19.15 |
| **Emotion** | 3.653 | 204.50 | .687 | 19.24 | 20.73 |
| **Pro-Social** | 3.756 | 141.00 | .175 | 22.58 | 17.55 |
| **Social** | 2.083 | 158.00 | .380 | 21.68 | 18.40 |
| **Social Behaviour** | 2.113 | 126.00 | .074 | 23.37 | 16.80 |
| **Tone** | 2.532 | 158.50 | .380 | 21.66 | 18.43 |
| **Word Count** | 1.958 | 236.00 | .204 | 17.58 | 22.30 |

## Discussion

The aim of the current study was to investigate, for the first time, whether AoC patients perceive and report social interactions differently compared to HVs. Using automatic linguistic processing tools and inferential statistics, the hypothesis was not supported: findings revealed no significant differences in the social and emotional content used to describe social interactions. Bayesian analyses could confirm that there was three times more evidence in favour of the null hypothesis (suggesting moderate evidence of a real null effect) for ‘emotion’ and ‘pro-social’ word use.

These results are surprising in light of the admittedly small but growing collection of studies investigating the impact of AoC on patients social functioning, finding that patients often miss social events, have a shrinking social network, and report decreased pleasure in social interactions compared to HVs (13,14). Consequently, it was hypothesised that AoC patients would describe social interactions differently compared to HVs. Specifically, that they would use fewer words associated with social behaviour according to LIWC categories (pro-social, social, social behaviour). Indeed, Bayesian analysis confirmed that for the ‘pro-social’ category there was more evidence in favour of a genuine null effect. However, it is acknowledged that the remaining categories were identified as insensitive and thus suggest that the current study may be underpowered. Further research is required to investigate this finding.

All experimental psychological studies to date have instead examined emotional processing in AoC, finding that AoC patients struggle to identify other’s emotions (14–16), report lower trait empathy (15) and higher alexithymia (12). Thus, it is also surprising that there were no differences in the use of emotion related words between AoC participants and HVs in the current study. Again, Bayesian analysis confirmed that for the ‘emotion’ category there was more evidence in favour of a genuine null effect.

Taken together, the surprising absence of significant results may be a positive indication that AoC patients do not struggle to identify or describe social interactions. There are, however, several alternative explanations. First, it may be that the task used in the current study was not the right task to capture the difficulties identified in previous research (see below). Second, it may be a result of selection bias. Previous research indicates that the extent of hypothalamic involvement of craniopharyngiomas strongly relates to patient’s physical and psychological functioning (1,3,8,25). Therefore, the current study may mask potential differences if the study happened to recruit individuals with low levels of hypothalamic involvement. Finally, and relatedly, the extent of hypothalamic or pituitary damage may impact the production and secretion of the hormone oxytocin which is known to play a vital role in both emotion processing (26,27) and social behaviour (28). Again, therefore, the current study may mask potential differences if AoC participants did not exhibit an oxytocin deficit. While it was beyond the scope of the current study to measure oxytocin, nearly half of AoC participants reported they had a diagnosis of Diabetes Insipidus which is characterised by a deficit in arginine vasopressin, the sister peptide of oxytocin, which is synthesised and produced in the same way as oxytocin, and which previous research has demonstrated that patients with Diabetes Insipidus (as a result of neurosurgery) also present with an oxytocin deficit (15). Thus, it is plausible that the AoC sample may have contained at least some patients with low oxytocin. Nevertheless, further research is required to investigate the influence of *both* hypothalamic involvement and oxytocin deficiency in social functioning in AoC.

It should be noted, however, that although the current study cannot speak to hypothalamic involvement or oxytocin concentrations, the sample was generally representative of other clinical characteristics of AoC: all patients had undergone surgical resection of the tumour; 58% had undergone additional radiotherapy; 47% reported they had Diabetes Insipidus; 68% were on hormone replacement therapy, including 21% on growth hormone; and finally, 79% reported some amount of visual disturbance as a result of their diagnosis or treatment, of which 21% said this was severe.

The current study has various strengths: i) recruiting verified AoC patients; ii) with matched HVs; iii) using an online experimental task enabled wider recruitment opportunities; iv) the sample size is comparable to, or larger than, all existing psychological studies (11,12,14–16); v) the study was pre-registered, and the data and code are freely available in line with open science practices. Nevertheless, some limitations are acknowledged. The use of automatic linguistic processing, while useful in many ways, may not be sensitive enough or the right tool for the current aim. While the LIWC dictionaries were screened for relevant categories (and were pre-registered) admittedly, words such as ‘you’, ‘he’, ‘she’ are perhaps not the words one might anticipate as relevant for a ‘social’ category. Therefore, the absence of significant group difference may not be surprising given the specific words LIWC searches for within certain categories. Relatedly, although the stimuli used are tightly controlled, previously validated and ecologically valid, the use of 4-second videos may have masked group differences. To address these two limitations, future research could use longer clips or in-person interactions with more in-depth coding analysis. While the online nature of the study facilitated recruitment, it may also have hindered the quality of data collection, with in-person verbal descriptions potentially revealing more information.

Additionally, although the sample size is comparable to previous studies, Bayesian analyses suggest that for *some* of the categories the data were insensitive, suggesting the study may be underpowered. However, it is i) extremely difficult to recruit participants with rare medical conditions, and ii) extremely difficult to determine adequate sample sizes where no previous evidence exists. It is likely that the effect sizes for social perception vary from the small collection of studies reporting experimental data on emotion processing in AoC, making power calculations unreliable. Thus, the current study serves as an important first step in this new area of investigation.

In conclusion, the present study was the first experimental study to investigate whether AoC patients perceive and report social interactions differently compared to HVs. Bayesian analysis confirmed there were no differences in the use of pro-social or emotion words between AoC participants and HVs. Further research is required to investigate and explain, and ultimately to intervene and improve, social and emotional functioning in AoC.

## Data Availability

The data and code are available online (https://osf.io/ux2kp/?view_only=47b518305d0e4cceb156f95a14f6ae80).

## Acknowledgements

The authors thank the various charities and platforms that assisted in recruitment and the participants for taking part in the research.

## Funding

This study was supported by the Essex ESNEFT Psychological Research Unit.

## Disclosure

The authors declare no conflicts of interest.

## Notes

### Competing Interest Statement

The authors have declared no competing interest.

### Author Declarations

The study was approved by the Ethics Committee at the University of Essex.

